# Functionalized TiO_2_ nanotube-based Electrochemical Biosensor for Rapid Detection of SARS-CoV-2

**DOI:** 10.1101/2020.09.07.20190173

**Authors:** B.S. Vadlamani, T. Uppal, S.C. Verma, M. Misra

## Abstract

The COronaVIrus Disease (COVID-19) is a newly emerging viral disease caused by severe acute respiratory syndrome coronavirus 2 (SARS-CoV-2). Rapid increase in the number of COVID-19 cases worldwide led the WHO declare pandemic within a few month after the first case of infection. Due to the lack of a prophylactic measure to control the virus infection and spread, early diagnosis and quarantining of infected as well as the asymptomatic individuals are necessary for the containment of this pandemic. However, the current methods for SARS-CoV-2 diagnosis are expensive and time consuming although some promising and inexpensive technologies are coming out for emergency use. In this work, we report the synthesis of a cheap yet highly sensitive cobalt-functionalized TiO_2_ nanotubes (Co-TNTs)-based electrochemical biosensor and its efficacy for rapid detection of spike glycoprotein of SARS-CoV-2 by examining S-RBD protein as the reference material. A simple, low-cost, and one-step electrochemical anodization route was used to synthesize TNTs, followed by an incipient wetting method for cobalt functionalization of the TNTs platform, which is connected to a potentiostat for data collection. This sensor specifically detected the S-RBD protein of SARS-CoV-2 even at very low concentration (range of 14 nM to 1400 nM). Additionally, our sensor showed a linear response in the detection of viral protein with concentration range. In summary, our Co-TNT sensor is highly effective in detecting SARS-CoV-2 S-RBD protein in approximately 30 seconds, which can be explored for developing a point of care diagnostics for rapid detection of SARS-CoV-2 in nasal secretions and saliva samples.

**AUTHOR SUMMARY:** SARS-COV-2 is currently a global pandemic on a scale that has not been experienced since the Spanish flu of 1918. One of the reasons why this pandemic virus has spread so quickly is because many infected individuals with SARS-CoV-2 remain asymptomatic and involuntarily transmit the virus before they come down with the symptoms. Therefore, uniform surveillance and quarantining of infected as well as the asymptomatic individuals could provide an effective measure to contain the spread of SARS-CoV-2. However, the current methods for SARS-CoV-2 diagnosis are expensive and time consuming although some inexpensive technologies are getting approvals for emergency use. Our manuscript reports the synthesis of a cheap yet highly sensitive cobalt-functionalized TiO_2_ nanotubes (Co-TNTs)-based electrochemical biosensor for rapid detection of spike glycoprotein of SARS-CoV-2. Our sensor is synthesized through one-step electrochemical anodization route, followed by an incipient wetting method for cobalt functionalization of TNTs platform. The readout of this sensor is an electrochemical signal collected through a potentiostat, which can be adopted for use through smartphone applications and the development of a point of care diagnostics for COVID-19.

## INTRODUCTION

The current outbreak of novel coronavirus (nCoV-2019 or SARS-CoV-2), was first detected in Wuhan, China in December 2019, but quickly spread to other parts of China as well as to the entire world causing pandemic [1]. According to the WHO, as of 16^th^ August 2020, around 21,294,845 people are infected, and 761,779 people have died due to SARS-CoV-2 infection [2]. SARS-CoV-2 infection causes a variety of symptoms including fever, cough and respiratory distress, which are collectively called as coronavirus disease or COVID-19 [3]. The transmission of SARS-CoV-2 primarily occurs from person to person through close contact or via small droplets produced during coughing, sneezing, and talking [4][5]. The incubation period for SARS-CoV-2 is around 2-7 days, with no noticeable symptoms; however, the viral transmission from an infected person to a non-infected person is still possible during this asymptomatic period [6]. Under the current scenario, with no vaccines in the market, global lockdown regulations are in place in order to minimize the viral spread. Consequently, this pandemic has caused a severe socio-economic impact on the world economy and raised fears of a global recession [7]. Currently, the real-time reverse-transcriptase polymerase chain reaction (RT-PCR) technique is the most common and reliable laboratory testing method for qualitative/quantitative SARS-CoV-2 detection [8][9] followed by serum virus neutralization assay (SVNA) for the determination of antibody neutralization [10] and enzyme-linked immunoassays (ELISA) for the detection of antibody against SARS-CoV-2 [11]. However, the major limitations of these laboratory based diagnostic tests is the invasive nature of the tests that often require trained personal for nasopharyngeal sample collection, along with the requirement of highly sophisticated machines, cross-reactivity with other viruses, and longer duration of testing. In order to contain the viral spread, surveillance of even asymptomatic individuals are needed, which is feasible only after the development of a simple, portable and rapid point-of-use sensor for the detection of SARS-CoV-2.

SARS-CoV-2 has positive-sense, single-stranded RNA (∼30K bp) genome with 14 ORFs that encode for structural, replication and non-structural proteins [12]. Similar to its genetic cousin, human SARS-CoV, SARS-CoV-2 consists of four structural proteins viz. spike (S), envelope (E), membrane (M), and nucleocapsid (N). Coronaviruses are named for the crown like spike glycoprotein, S (composed of two subunits: the S1 subunit and S2 subunit) on the surface/envelop [13]. The S1 subunit of the S protein consists of a receptor binding domain (RBD) that has a high binding affinity towards the host angiotensin-converting enzyme II (ACE2) receptor present on the human cells and the S2 subunit mediates virus-host cell fusion and entry [14]. Importantly, the S protein is highly immunogenic and induces immune response to produce neutralizing antibodies as well as T-cell responses in SARS-CoV-2 infected individuals [15]. Functionally, binding of the S-RBD to the hACE2 receptor is a crucial for the entry of SARS-CoV-2 into the human cells. Infringingly, SARS-CoV-2 S-RBD shares only 70% sequence identity with SARS-CoV S-RBD, which has been evaluated for vaccines and therapeutic drug development [16]. Hence, the S-RBD of SARS-CoV-2 are excellent target for diagnostic and therapeutic intervenstions.

Electrochemical biosensors are advantageous for sensing biomolecules because of their ability to detect biomarkers with accuracy, specificity and high sensitivity [17]. Electrochemical biosensors have been successfully used in medical diagnostics for the detection of viruses such as Middle East respiratory syndrome coronavirus (MERS-CoV),[18] human enterovirus 71 (EV71) [19], human influenza A virus H9N2 [20], avian influenza virus (AIV) H5N1 [21]. Lahyquah *et al*.[18] used an array of carbon electrodes modified with gold nanoparticles for the detection of MERS-CoV. Very recently, a biosensor using gold nanoparticle decorated FTO glass immobilized with nCovid-19 monoclonal antibody was reported for the detection of SARS-CoV-2 [22]. The functionality of the electrochemical biosensor can be further improved by nanostructuring the electrode as it increases the electrochemical reaction rate due to an increased electrode surface area to volume ratio, thereby increasing the electrode surface area to analyte fluid volume. In the work by Chin *et al*. on the encephalitis virus, it was found that nanostructuring of carbon electrodes with carbon nanoparticles increased the current response by 63% due to an enhanced electron charge transfer kinetics [23]. Similarly, we have reported that Co functionalized TiO_2_ nanotubes (Ni-TNTs) with higher surface-to-volume ratio can detect the biomarkers associated with tuberculosis [24][25]. The proposed sensing mechanism involves the formation of a complex between Co and the biomarker at specific bias voltage, due to the reduction of Co ions and oxidation of biomarker. Similarly, we hypothesized that S-RBD or SARS-CoV-2 can be detected through complexing of functionalized nanoparticles with the S-RBD protein and a schematic of viral detection directly from patient sample as shown in figure 1.

**Figure 1.**
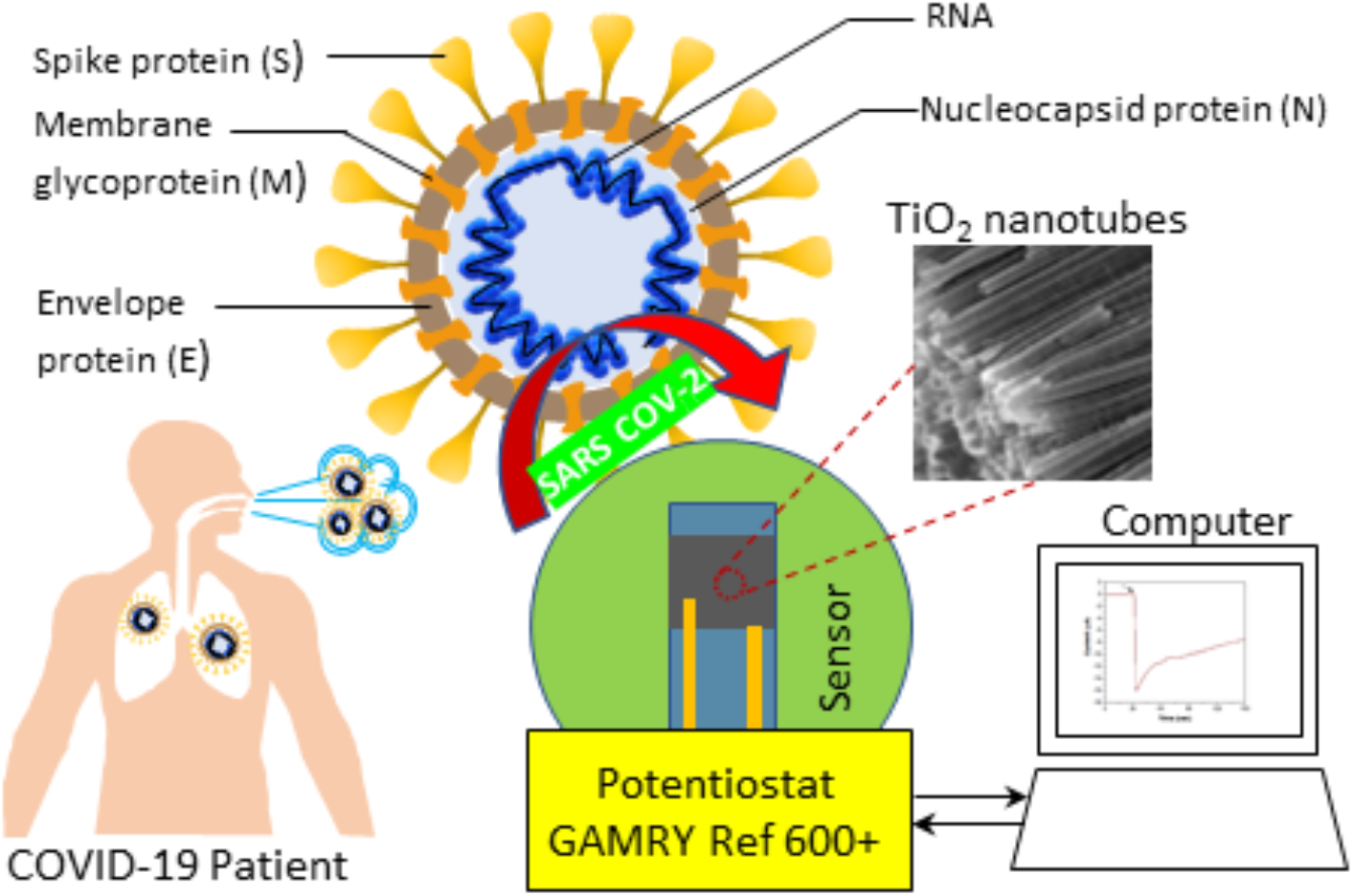
Schematic of Co functionalized TNT based sensing platform for the detection of SARS-CoV-2.

In the current work, we have determined the potential of Co-functionalized TiO_2_ nanotubes (Co-TNTs) for the electrochemical detection of S-RBD protein of SARS-CoV-2. TNTs were synthesized by simple, cost-effective, one-step electrochemical anodization route, and Co functionalization was carried out using the incipient wetting method. Our data showed that cobalt functionalized TNTs could selectively detect the S-RBD protein of SARS-CoV-2 using the amperometry electrochemical technique in ∼ 30 secs.

## MATERIALS AND METHODS

### Synthesis of TNTs

TNTs were synthesized by electrochemical anodization of Ti sheet. Ti sheet of size 1.5 cm x 1.5 cm, with a tab 2 mm in width, was cut out of G1 grade Ti sheet (thickness 0.01016 mm). One side of the coupon was polished with 600 grit polishing paper for 4 min to remove any surface metal oxide layer. The coupon was ultrasonicated in 1:1 solution of ethanol and acetone for 2 min. The unpolished side was masked with Kapton tape to avoid any exposure to electrolyte during anodization. The electrochemical anodization was performed in a standard two-electrode configuration, using Ti foil as a working electrode and platinum foil as a counter electrode with a 3 cm gap between them. The anodization was carried out using an electrolyte of composition 96.5 ml (CH_2_OH)_2_, 3 ml DI H_2_O, and 0.505 g NH_4_F in a Teflon beaker. The electrolyte was maintained at a subzero temperature and continuously stirred using a magnetic stirrer at a speed of 140 rpm. The anodization was carried by maintaining a constant voltage of 30 V across both the electrodes for 50 min. After anodization, the sample was rinsed in DI H_2_O and baked in an oven at 120°C for 4 hrs. The Kapton tape was removed from the sample after baking, and the sample was annealed in a tube furnace at 500°C for 3 h in a continuous flow of oxygen.

### Synthesis of Co functionalized TNTs

The annealed TNTs obtained from the furnace were functionalized with cobalt using an incipient wetting method, i.e. a wet ion exchange process. The same side of the sample that was masked earlier was again masked with Kapton tape. The sample was ultrasonicated in a solution containing 2.306 g of CoCl_2_.6H_2_O in 20 ml ethanol for 35 min. The sample was baked in an oven at 120°C for 4 hrs to obtain cobalt functionalized TNTs.

### SEM Characterization

The morphology of the TNTs and Co-TNTs were examined using Dual Beam Scanning Electron Microscopy (SEM, ThermoFisher Scientific). The cobalt content in the Co-TNTs sample was analyzed using the EDS detector attached to SEM. The SEM micrographs were analyzed using ImageJ software.

### Synthesis and Purification of SARS-CoV-2 S-RBD protein

The pCAGGS vector containing SARS-CoV-2 Wuhan-Hu-1 Spike Glycoprotein Receptor Binding Domain (RBD) with a C-terminal hexa-histidine tag was obtained from BEI Resources (NIAID, NIH, NR-52309). His_6_-tagged S-RBD containing pCAGGS plasmid was expressed in HEK293T (human embryonic kidney) cells obtained from ATCC (American type culture collection) and maintained in Dulbecco’s modified Eagle medium (DMEM) supplemented with 10% fetal bovine serum (FBS, Atlanta Biologicals), 2 mM L-glutamine, 25 U/mL penicillin, and 25 μg/mL streptomycin. Cells were grown at 37° C in a humidified chamber supplemented with 5% CO_2_. For His_6_-tagged S-RBD protein generation, HEK293T cells were transfected with recombinant plasmid using Neon transfection system (Thermo Scientific) according to the manufacturer’s instructions. Supernatants from transfected cells were harvested on day 3 post-transfection and the cell debris was removed by centrifugation (4,000 rpm, 20 min at 4° C). Supernatants were then incubated with 1mL of Ni-NTA Agarose (Qiagen) for every 10 mL of supernatant, for 2 hours at 4°C with rotation. For S-RBD purification, gravity flow columns were used to load the NI-NTA agarose bound His_6_-tagged Spike-RBD protein, followed by washing with wash buffer (20mM sodium phosphate, 500 mM NaCl, 8 M urea, 20 mM imidazole, pH 6.0) and eluting with elution buffer (20mM sodium phosphate, 500mM NaCl, 8M urea, 200mM imidazole, pH 4.0). Eluted protein was concentrated using protein concentrators (Thermo Scientific, 87748 and 87772), quantified using Bradford assay and Nanodrop (Thermo Scientific) and further analyzed by SDS-PAGE.

### Electrochemical Characterization

The electrochemical sensing of S-RBD protein was carried out using a custom-built Co-TNT packaged printed circuit board setup. The sensor response was measured with the help of Gamry reference 600+ Potentiostat attached to the printed circuit board. The schematic of the whole sensing set up along with the detection methodology is shown in Figure 1. The sensor response with various S-RBD protein concentrations was determined using the amperometry technique, at a bias voltage of −0.8 V. The bias voltage was determined by conducting the cyclic voltammetry experiments in the voltage window −2 V to +2 V. All the experiments were carried out at room temperature.

## RESULTS and DISCUSSION

### Co-TNT showed characterstics nanotube formation

The Scanning Electron Microscopy (SEM) micrographs of the TNTs, prepared by electrochemical anodization, are shown in Figure 2a. The inset shows the side view of the TNTs (Figure 2a). The outer diameter and wall thickness of TNTs were ∼60 nm and ∼10 nm, respectively. The average length of TNTs was found to be ∼1.1 µm. In our earlier work, TNTs synthesized under similar conditions were found to show the crystalline anatase phase predominantly [25].

**Figure 2.**
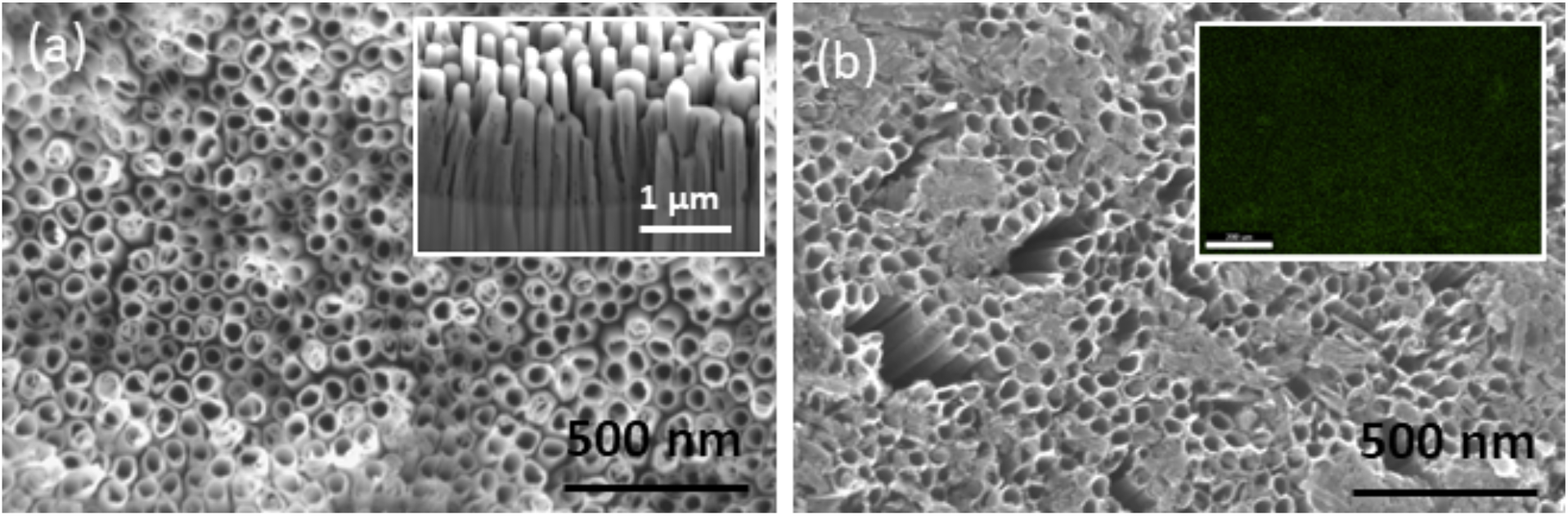
SEM micrographs of (a) TNTs post-annealing. Inset shows sidewalls of TNTs and (b) Co-functionalized TNTs showing the Co(OH)_2_ precipitate. Inset shows an EDS map of Co confirming its uniform distribution.

The surface morphology of the Co-TNTs examined under SEM is shown in Figure 2b. The SEM micrograph reveals the presence of precipitates on top of the TNT surface. EDS analysis confirmed the uniform distribution of Co on top of TNTs, and the Co content was found to be ∼ 4 wt %. We have previously shown that Co exists in Co^+2^ state in the form of Co(OH)_2_ on Co-TNTs [26]. Therefore, the morphology of TNTs can be visualized as having a very large surface area, uniformly decorated with Co^+2^ ions.

### S-RBD Protein showed specific monomeric and dimeric forms

The S-RBD protein, biomarker for SARS-CoV-2 detection, was characterized via SDS-PAGE (under denaturizing conditions). The RBD domain of spike glycoprotein comprises of amino acids 329-521, which is a ∼25 kDa protein with potential N-glycosylation sites. As shown in Figure 3a-b, the SDS-PAGE gel of His_6_-tagged S-RBD protein was either stained with SimplyBlue SafeStain (Figure 3a) or transferred to a nitrocellulose membrane and detected with 1 ug/mL of mouse anti-His monoclonal antibody (Figure 3b), followed by incubation with infrared-dye-tagged secondary IR-Dye680 antibody and scanning with an Odyssey infrared scanner. Specific bands were detected for SARS-CoV-2 S-RBD protein at approximately 35kDa and 70kDa, representative of the monomeric and dimeric forms of S-RBD protein, respectively (Fig. 3B). We detected the S-RBD at a slightly higher molecular weight (∼35kda) possibly because of post-translational modifications including glycosylation.

**Figure 3.**
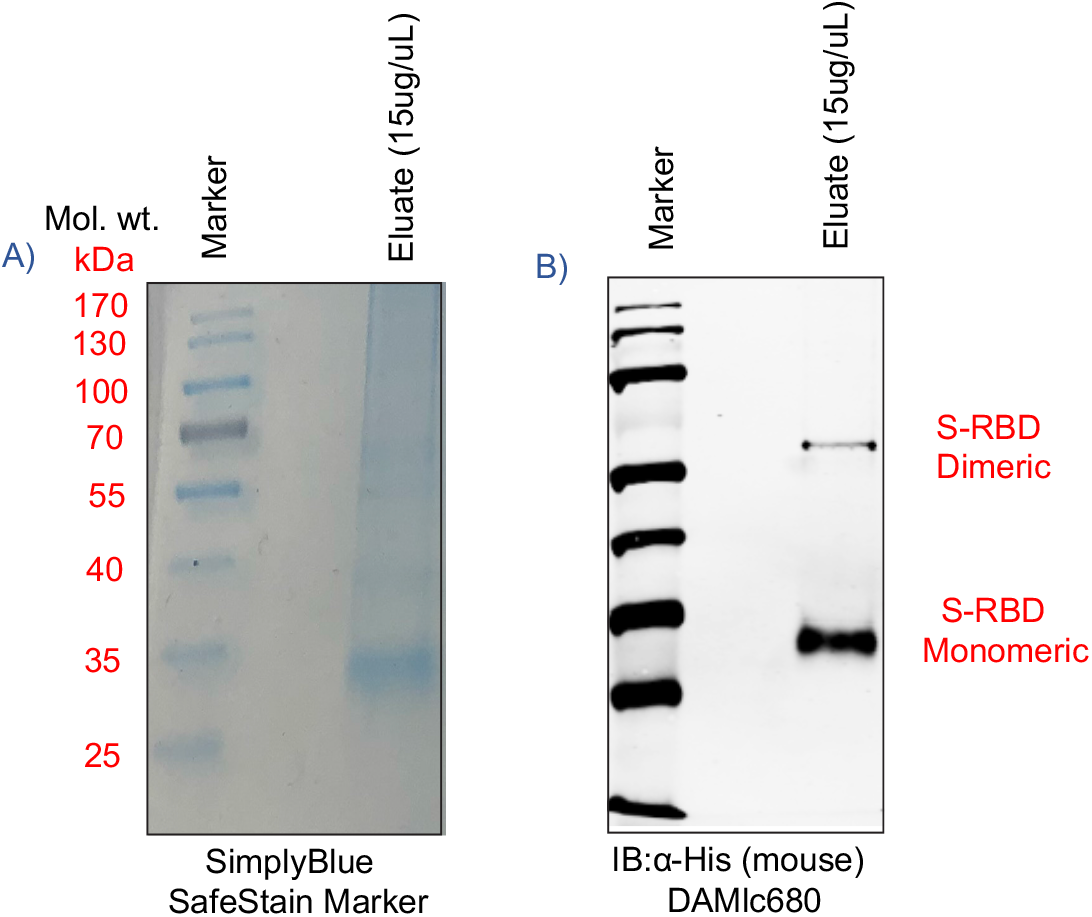
A) SimplyBlue SafeStained gel, and B) Western blot of His_6_-tagged S-RBD protein (purified under denaturing conditions) and probed with mouse anti-His monoclonal antibody. SARS-CoV-2 S-RBD characteristic bands were detected at 35kDa/monomeric form and 70kDa/dimeric form, respectively.

### S-RBD Protein was detected on Co-TNTs sensors

The ability of Co-TNT to sense S-RBD protein of SARS-CoV-2 was determined by performing an amperometry experiment at a bias voltage of −0.8 V. The amperometry curves obtained at various concentrations of protein are shown in Figure 4. The sensor was exposed to protein 30 sec after beginning of experiment (marked by an arrow). The sensor response current increases sharply and rapidly as the sensor was exposed to the protein. At a protein concentration of 1400 nM (nano molar), the peak sensor current output was found to be ∼0.74 µA (nano ampere). The peak current decreases to ∼0.45 µA at a protein concentration of 140 nM and further decreases to ∼0.23 µA at a protein concentration of 14 nM. The sensor detection time was ∼ 30 sec over the concentration range of 14 nM to 1400 nM. It is hypothesized that the rapid increase in sensor response current could be attributed to the electrochemically triggered unfolding of protein that exposes its interior [27][28][29] and subsequent complex formation between Co and the protein [30][31][32].

**Figure 4.**
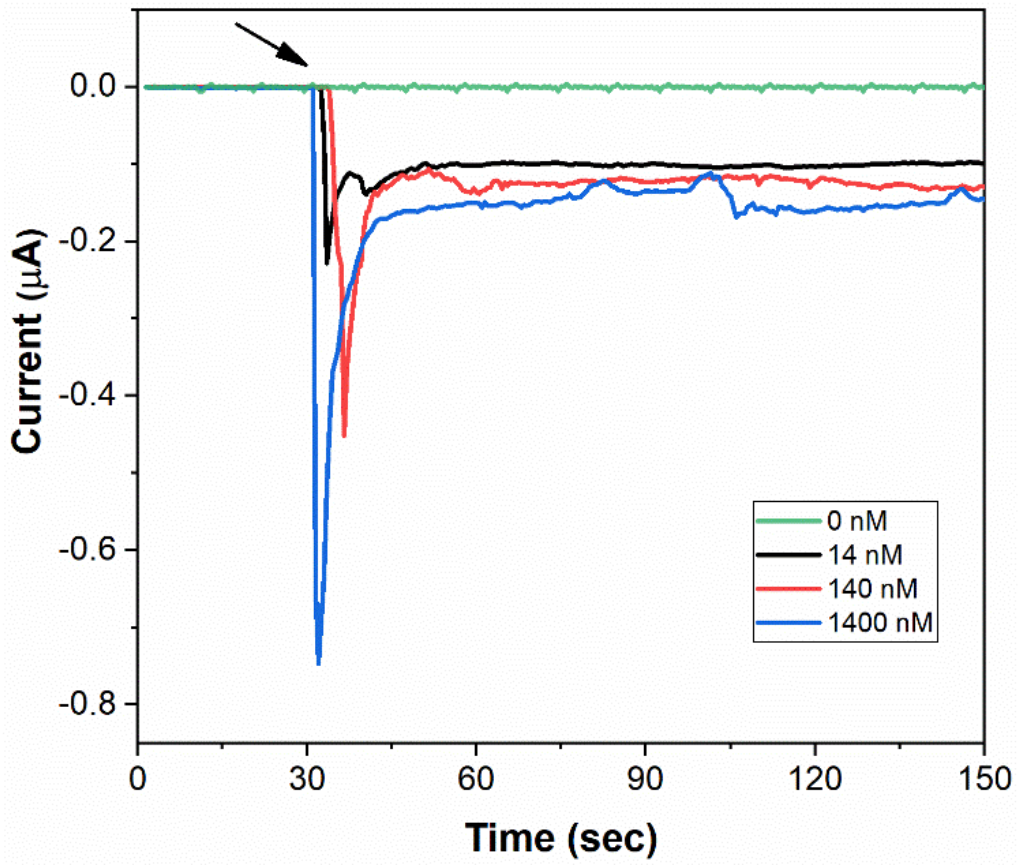
Amperometry response curves of Co-TNT sensor, at a bias voltage of −0.8 V, upon exposure to SARS-CoV-2 S-RBD protein of concentrations 0 nM (background), 14 nM, 140 nM, and 1400 nM.

The average sensor response time, which is defined as the time taken to reach the peak current, was found to be ∼ 2 sec. It is very short compared to our earlier studies on the sensor for Colorectal Cancer, where a sensor response time of ∼200 sec was documented [33]. The shorter sensor response time indicates higher kinetics of reaction between Co-TNT and the protein molecules.

### Sensor response measurement

The sensor response (SR) was calculated at various protein concentrations based on the following equation:

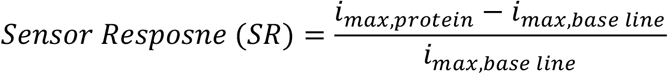

where *i*_*max,protein*_ is the maximum current obtained when sensor is exposed to SARS-CoV-2 S-RBD protein and *i*_*max,base line*_ is the maximum current obtained when sensor is not exposed to the protein. The value of *i*_*max,base line*_, which is the current obtained when sensor is not exposed to protein, was found to be ∼10 pA (Figure 4). The sensor responses measured at different protein concentrations are shown in Figure 5. The sensor response was found to increase with an increase in the concentration of protein. Moreover, the sensor response exhibited excellent linearity over the concentration range 14 to 1400 nM with a correlation coefficient of R^2^ = 0.99. The regressed linear calibration curve for sensor response was obtained as follows:

> *SR* = 0.266 ± 0.026) log(*C*) + (4.053 ± 0.059); R^2^ = 0.99

**Figure 5.**
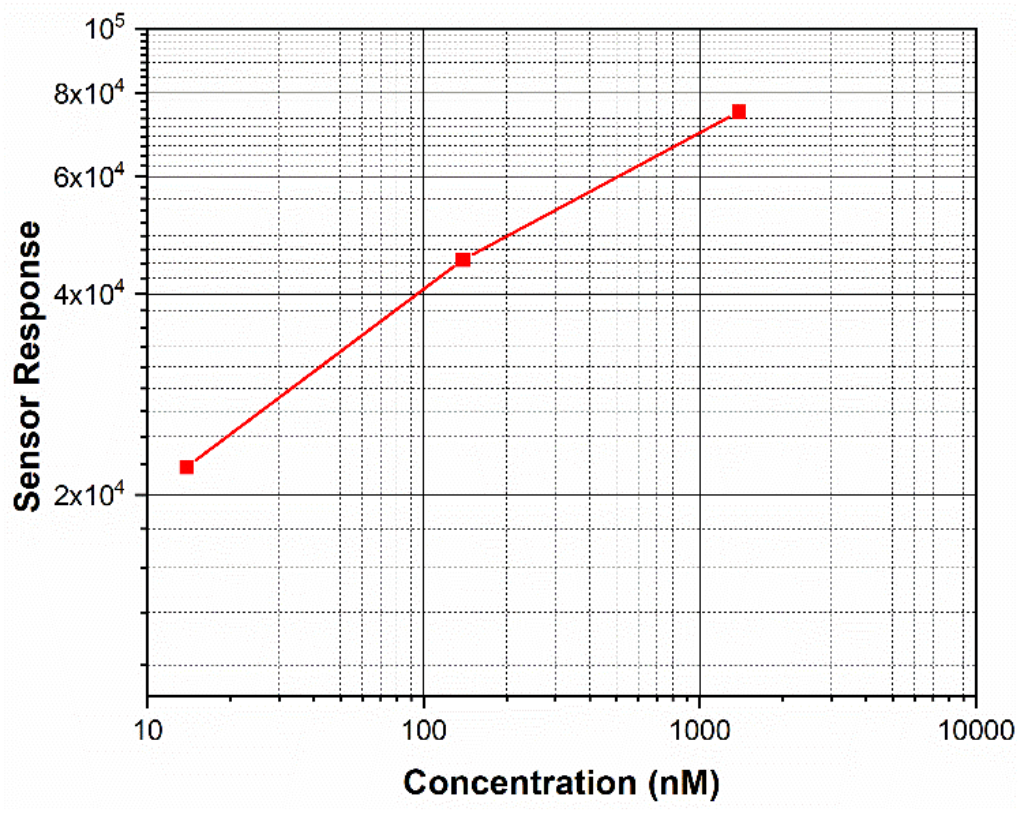
Plot showing the effect of SARS-CoV-2 S-RBD protein concentration on the variation of the sensor response for the Co-TNT sensor. The sensor response shows a linear region from 14 nM to 1400nM.

where SR is the sensor response, and C is the concentration of protein in nM. Using statistical analysis [34] the limit of detection for measurements made using sensor was determined to be 0.7 nM.

The limit of detection can be further improved by the use of (i) Co-TNT synthesized by in-situ anodization technique and (ii) Co-TNTs of even higher length. Previously, we found that Co-TNT synthesized by in-situ anodization with higher sensor sensitivity compared to Co-TNT synthesized by incipient wetting route towards the detection of tuberculosis biomarkers [25]. A higher sensor sensitivity corresponds to a better limit of detection and senstivity of quantitation. The increased sensitivity was attributed to the presence of Co(OH)_2_ precipitate sites in direct contact with parent TiO_2_ due to which direct conduction is possible. The sensor sensitivity can also be improved by using longer Co-TNTs as higher surface area results in a higher reaction rate; thereby, higher sensor response current can be obtained even at lower protein concentrations.

## CONCLUSIONS

In this study, we developed a Co-metal functionalized TNT as a sensing material for electrochemical detection of SARS-CoV-2 infection through the detection of the Receptor Binding Domain (RBD) of spike glycoprotein. We confirmed the biosensor’s potential for clinical application by analyzing the RBD of the Spike glycoprotein on our sensor. Amperometry electrochemical studies indicated that the sensor could detect the protein in the concentration range 14 nM to 1400 nM. The relationship between sensor response and protein concentration was found to be linear with the limit of detection as low as ∼0.7 nM levels. Importantly, our sensor detected SARS COV-2 S-RBD protein in a very short time (∼30 sec) confirming its implication in developing a rapid diagnostic assay. Thus, our report demonstrate the development of a simple, inexpensive, rapid and non-invasive diagnostic platform that has the potential of detecting SARS-CoV-2 on clinical specimens including nasal, nasopharyngeal swabs or saliva. Moreover, the developed approach has the potential for diagnosis of other respiratory viral diseases by identifying appropriate metallic elements to functionalize TNTs.

## Data Availability

All the data are provided in the manuscript.

## ACKNOWLEDGEMENTS

This work was supported by the departmental and institutional funds. The S-RBD expression vector was obtained through BEI Resources, NIAID, NIH: Vector pCAGGS Containing the SARS-CoV-2, Wuhan-Hu-1 Spike Glycoprotein Gene RBD with C-Terminal Hexa-Histidine Tag, NR-52309.

## AUTHOR CONTRIBUTION

BSV: formal analysis, methodology, writing – original draft preparation, writing – review and editing.

TU: methodology, writing – original draft preparation, writing – review and editing.

MM: conceptualization, methodology, project administration, funding, writing – review and editing.

SCV: conceptualization, methodology, project administration, funding, writing – review and editing.

## DECLARATIONS

### Approval

The Environmental and Biological Safety committee of the University of Nevada, Reno, approved methods and techniques used in this study. appropriate metallic elements to functionalize TNTs.

